# Assessment of Ankle Fractures using Deep Learning Algorithms and Convolutional Neural Network

**DOI:** 10.1101/2021.07.25.21261103

**Authors:** Soheil Ashkani-Esfahani, Reza Mojahed Yazdi, Rohan Bhimani, Gino M. Kerkhoffs, Mario Maas, Daniel Guss, Christopher W. DiGiovanni, Bart Lubberts

**Author notes:** Correspondence: Reza Mojahed-Yazdi, MSc, Address: Yawkey 3, 55 Fruit Street, Massachusetts General Hospital, Boston, 02114, MA. **Funding** The author(s) received no financial support for the research, authorship, and/or publication of this article. **Place of study:** This study was conducted in Foot and Ankle Research and Innovation Laboratory, Massachusetts General Hospital, Harvard Medical school, Boston, 02114, MA. **Author Contributions Statement:** All authors have participated in designing the study, data acquisition, analysis, and interpretation, as well as writing the draft, and critical revision of the paper. The final manuscript was approved by all the authors.

## Abstract

Early and accurate detection of ankle fractures is crucial for reducing future complications. Radiographs are the most abundant imaging techniques for assessing fractures. We believe deep learning (DL) methods, through adequately trained deep convolutional neural networks (DCNNs), can assess radiographic images fast and accurate without human intervention. Herein, we aimed to assess the performance of two different DCNNs in detecting ankle fractures using radiographs compared to the ground truth.

In this retrospective study, our DCNNs were trained using radiographs obtained from 1050 patients with ankle fracture and the same number of individuals with otherwise healthy ankles. Inception V3 and Renet50 pretrained models were used in our algorithms. Danis-Weber classification method was used. Out of 1050, 72 individuals were labeled as occult fractures as they were not detected in the primary radiographic assessment. Using single-view radiographs was compared with 3-views (anteroposterior, mortise, lateral) for training the DCNNs.

Our DCNNs showed a better performance using 3-views images versus single-view based on greater values for accuracy, F-score, and area under the curve (AUC). The sensitivity and specificity in detection of ankle fractures using 3-views were 97.5% and 93.9% using Resnet50 compared to 98.7% and 98.6 using inception V3, respectively. Resnet50 missed 3 occult fractures while Inception V3 missed only one case.

**Clinical Significance:** The performance of our DCNNs showed a promising potential that can be considered in developing the currently used image interpretation programs or as a separate assistant to the clinicians to detect ankle fractures faster and more precisely.

**Level of evidence:** **III**

## Introduction

Ankle injuries account for 5 million emergency department (ED) visits in the United States, 15% of which are fractures ^1^. Fractures of the ankle joint account for 9% of all fractures and 36% of those involving the lower limbs, making them one the most common fractures treated in the orthopedic settings ^1^. When inadequately diagnosed or left untreated, ankle fractures can be debilitating and result in significant long-term morbidities ^2-4^. The diagnosis of ankle fractures usually relies on clinical assessment supported by imaging techniques ^5^. Despite the increasing availability of modern imaging modalities including computed tomography (CT) and magnetic resonance imaging (MRI), the use of conventional radiographs has remained fundamental in the primary assessment of these patients. Radiographs have been the mainstay of initial assessment due to ease of access, time to perform, cost, and low radiation, particularly in resource-limited settings ^6^. In spite of these inherent benefits, about 23% of the ankle fractures are missed on the initial radiographic evaluation ^7^. Enhancing radiographic imaging machines with automated interpretation systems can lead to a considerable improvement in the accuracy and cost-efficiency of the present diagnostic protocols for ankle fractures.

Several studies have demonstrated the utility of machine learning for the detection of orthopedic conditions in musculoskeletal images ^8-12^. Deep learning (DL), a subset of machine learning, is quickly progressing and becoming a powerful tool in interpreting clinical images utilizing deep convolutional neural networks (DCNNs) ^13^. Rather than requiring manual feature assessment that is currently used for interpretation of clinical images, DCNNs allow the machine to use native image inputs such as pixels, Hounsfield differences, distances, brightness, contrast, etc. for assessing the images and comparisons ^13^. The more a DCNN is trained with a greater amount of data, the more accurate it performs. An alternative for providing a huge database to train a comprehensive neural network is to use a pre-trained DCNN. By using these DCNNs one only needs to retrain the last few layers of the network and the other layers will remain relatively unchanged. This can lead to even greater performance of the DL algorithm in the detection of musculoskeletal pathologies ^14^.

The purposes of this study were to evaluate the performance of DL algorithms and DCNNs in the detection and annotation of ankle fractures using radiographs, particularly occult fractures that were missed by the primary care clinicians. Can a DL-based algorithm detect ankle fractures with the same accuracy, if not better, compared to the human interpreter? Do 3-views radiographic assessment lead to a higher accuracy compared to the single-view and should we still obtain three views? Do DL-based methods have to potential to be used as an assistant, if not a substitute, to improve the accuracy and the speed of diagnosis for ankle fractures?

## Materials and Methods

### Study design and population

This research was a retrospective case-control study. The protocol of this study was approved by the Institutional Review Board (IRB; no. 2015P000464) and the requirement for informed consent was waived due to the retrospective design of the study and the use of deidentified patients’ data according to Health Insurance Portability and Accountability Act of 1996 (HIPAA) compliance.

Through a medical record search using the Research Patient Data Registry (RPDR) system from July 2015 to March 2020, 4930 patients who had undergone ankle X-ray radiographic evaluation, irrespective of weightbearing or non-weightbearing, were screened. From this population, 1660 patients with approved ankle fractures on X-rays or on a following CT scan examination were identified. The images were reviewed by a board-certified radiologist and orthopaedic surgeon to make sure the findings were concordant with the reports. Ankle fracture was defined as any bone fracture in the distal tibia, distal fibula, and body of the talus ^15^.

The inclusion criteria for the patient group were 1) Age ≥ 18 years, 2) detection of ankle fractures based on reports from X-ray or CT scan imaging ^16^, and 3) having 3-view -lateral, mortise, anteroposterior (AP)- X-ray radiographs. The exclusion criteria were 1) presence of hardware in the ankle bones due to previous operation, 2) having less than 3-view X-ray radiographs, 3) having any artifacts on the images and lesions other than fractures that change the normal texture of the bone including mal- or non-union after a previously detected fracture, cysts, masses, etc. Eventually, 1050 patients were included in this study. An equal-sized control group was selected comprised of 1050 individuals with healthy ankles without a history of ankle fracture based on radiological reports and examination notes. Among the patient group, in 72 individuals, the ankle fracture was not detected on initial X-rays. Instead, these fractures were diagnosed during further radiological assessments using additional X-rays and/or CT scans. We classified the fractures into three groups according to the Danis-Weber classification system ^17^. We selected the Danis-Weber classification due to its higher reproducibility compared to other classification systems ^17^. Briefly, fractures below the level of the syndesmosis were labeled as Weber A, at the level of the syndesmosis as Weber B, and above the level of the syndesmosis as Weber C. Images that could not be classified according to the Danis-Weber system- i.e. isolated fractures of the tibia or the body of talus, were labeled as “other”. Moreover, the presence or absence of concomitant fractures in the tibia and the talus were also assessed. All X-rays were extracted, anonymized, and numbered. Table-1 shows the demographic characteristics of the patient and control group as well as the distribution of the fractures in different Danis-Weber classes.

### Data preparation

In order to create the data stacks for the machine learning protocol, the 3-view images of each individual were labeled anteroposterior, mortise, and lateral, accordingly. The set of images were then inserted into a folder labeled as patient or control in the format of Digital Imaging and Communication in Medicine (DICOM) files. The radiographs were obtained using various radiography systems provided by different vendors. Thus, they differed regarding the properties of the images including the quality, brightness, and resolution. However, other than anonymization, we performed no additional pre-processing on the images including pixel value extraction, rescaling, and resizing. Our dataset was randomly split into train, validation, and final test subsets with a 60:20:20 split ratio. We utilized flipping and rotation (10° clockwise and counterclockwise) for data augmentation to increase generalization and enlarge the size of the available data set ^18^. Data augmentation leads to leveraging the data by applying minor changes to the primary dataset. This will increase the confidence in the results and validity of the algorithm as well as prohibiting overfitting.

### Machine learning protocol

Deep learning algorithms, as a subset of machine learning, are used to create deep convolutional neural networks (DCNN) by embedding multiple hierarchical hidden layers between input and output layers. The algorithm designs the feature extraction tool to learn and retain the underlying architectures in each class of data and uses them for mapping the final feature or outcome. To develop our DCNN and receive a better performance, we adopted the transfer learning technique. Transfer learning lets our algorithm use a DCNN architecture that has been pretrained for another relatively similar purpose- i.e. detecting lung abnormalities, to be used for our purpose which was differentiating healthy ankles from those with any fractures ^19^.

In this study we constructed different DCNNS based on two models, InceptionV3 and Resnet 50, to extract the features of radiographic images and create a feature map for the initial layers of the DCNN as outlined by the literature ^20, 21^. These models were pretrained with a huge database comprised of everyday images on ImageNet® (http://www.image-net.org) ^22^.

To construct our DCNN models, first, we designed a single-input architecture to incorporate single-view images. We selected the AP-view image as the most abundant image used by the clinicians to be used in this model. On the other hand, we designed two other architectures each receiving 3-view images of AP, mortise, and lateral, simultaneously. This 3-input structure passes each image through unique DCNNs followed by a global average pooling layer. Then, outputs of the previous step merge together using a concatenation layer followed by a single-unit dense layer. The sigmoid function is used to generate probability output and find the class values ^20, 21^. In our algorithms, we did not freeze the layer’s weight which was incorporated in the models that were pretrained on ImageNet®, instead, we provided freedom to all layers including the top and the bottom ones to be further developed and update their weights during the training process. During the training phase, the RMSProp method was employed to optimize the variables ^23^. The learning rate of *ι* = 0.001 and a batch size of n=256 was selected. The maximum number of epochs was limited to 200. We monitored the convergence of the model training using binary cross-entropy loss and the models were considered converged when the loss values reached the plateaued curve and showed no further decrease ^9, 24^. The early stopping technique is utilized to compare the performance of the model in different epochs and terminate the training process earlier if the model does not show any more capacity to decrease the test loss after 20 consecutive epochs ^25^. In other words, for example, if the model obtains the test loss value of 0.5 in the 10th epoch and cannot reduce this loss before the 30th epoch, the learning process will be ended. The network architecture was implemented using TensorFlow™ (Version 2.0, Google Inc., CA). All models were trained on Tesla K80 GPU with 12GB GGDDR5 RAM and 2496 Cuda cores. The architecture of our DCNNs is demonstrated in Figure 1. The accuracies of the algorithms based on the number of epochs are also shown in Figure 2.

**Figure 1.**
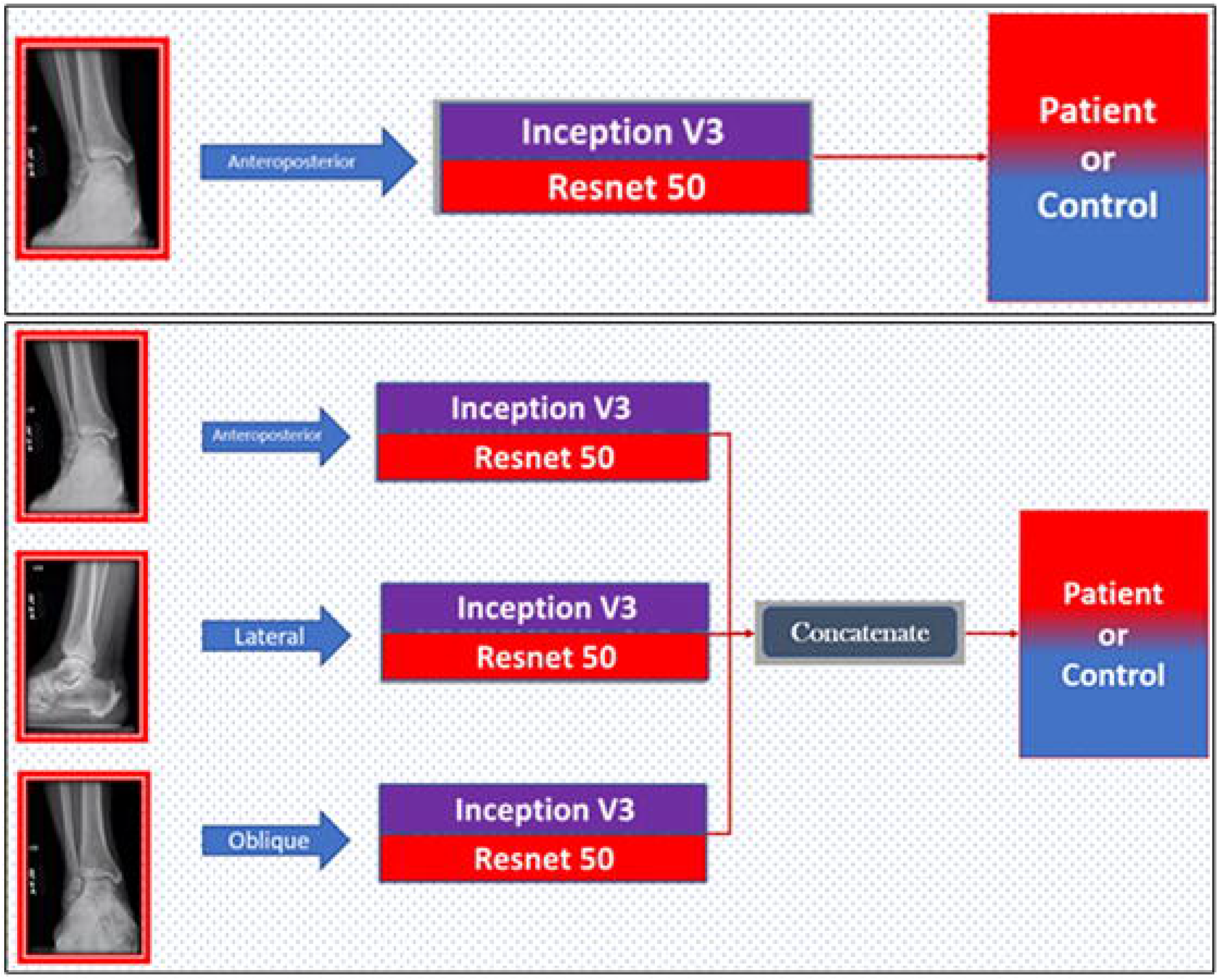
The architecture of the deep convolutional neural networks based on single-view and three-view radiographs using Inception V3 or Resnet 50 pretrained algorithms.

**Figure 2.**
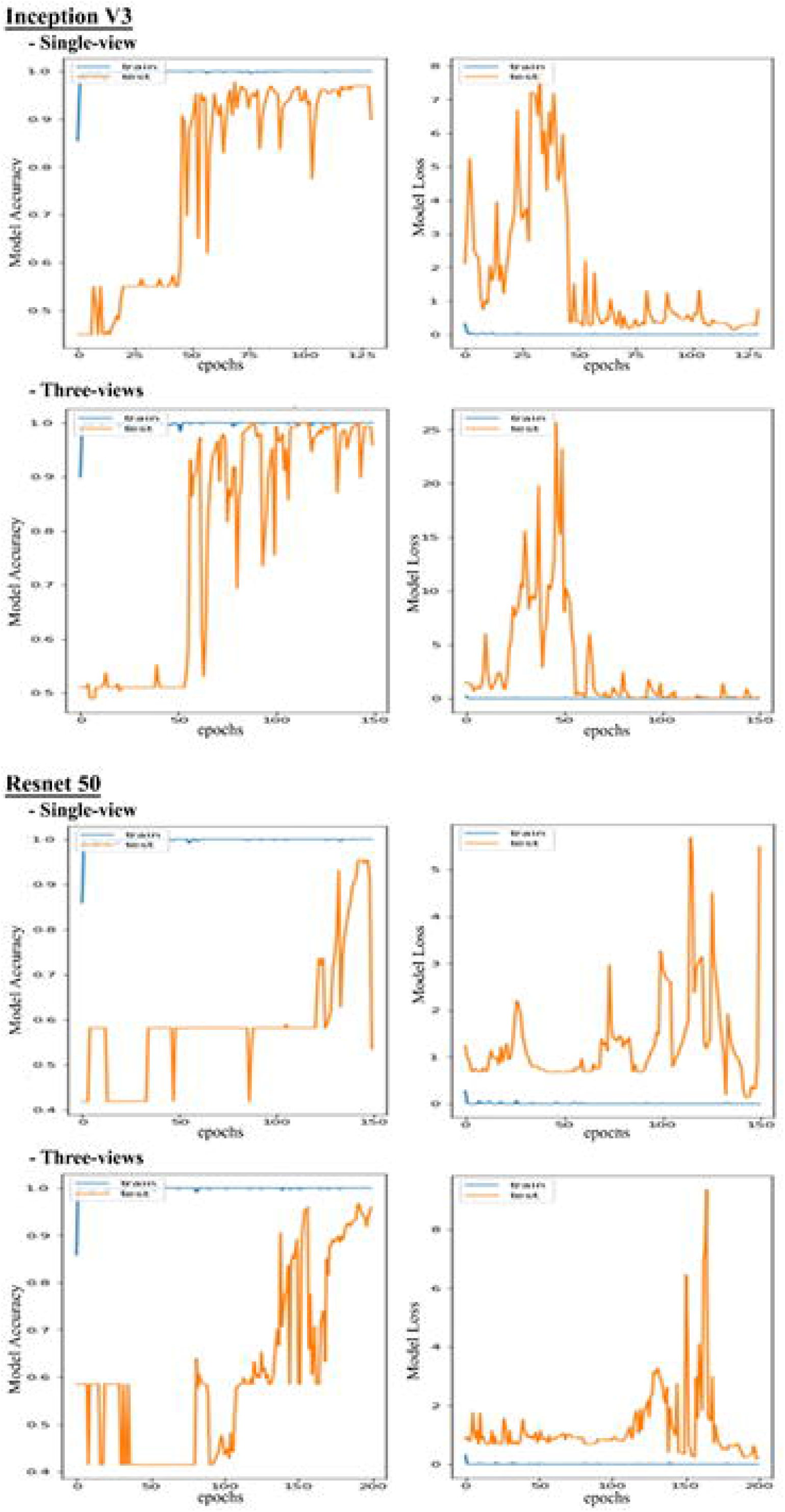
Accuracy and loss rates per epoch for each deep convolutional neural network based on single-view and three-view radiographs.

### Statistical analysis

To determine differences in demographics between the patients and the controls, independent samples t-test was used to compare values for age and body mass index (BMI); Chi-square was used to compare the frequency of each gender and also the frequency of fracture classifications. Data are demonstrated as mean and standard deviation (SD), or percentage. In order to exhibit the performance of the machine learning algorithms, sensitivity, specificity, positive predictive value (PPV), negative predictive value (NPV), area under the curve (AUC), accuracy, and F-score were reported. The AUC indicates the accuracy of a quantitative diagnostic test. A test with perfect accuracy has an AUC of 1 and an AUC of 0.5 indicates no discrimination. F-score is an indicator of accuracy which is commonly used for machine learning algorithms ^26^. For calculating the F-score we obtained the following formula: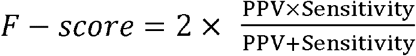. In this study p-vale < 0.05 was considered as statistically significant. Data analyses were performed using Stata 13.0 (StataCorp LP, College Station, TX).

## Results

Demographic data of the patients and the controls are shown in Table 1. There were no differences between the two groups regarding age, gender, and BMI. The proportions of ankle fractures based on Danis-Weber classification are demonstrated in Table 2. Among the fractures that were missed during the initial X-ray radiographic assessment (72/1050 total, 7%), 36/72 (50%) was a Weber A fracture, 24/72 (33%)were avulsion fractures of the talus body, 16% (11/72) were avulsion fractures of the medial malleolus, and one (1%) fracture was located in the proximal part of the fibula (Weber C).

**Table 1.** Baseline characteristics of the patients and the controls.

| **Group** | **Patient group** |  | **Control group** |  | **p-value** |
| --- | --- | --- | --- | --- | --- |
| **Gender** | M | 61% (643/1050) | M | 56% (592/1050) | 0.11 |
|  | F | 39% (407/1050) | F | 44% (458/1050) |  |
| **Age**<br>**(mean ± SD)** | 44.1±17.1 | | 38.9 ± 19.5 | | 0.28 |
| **BMI**<br>**(mean ± SD)** | 31.7±9.8 | | 28.1±11.2 | | 0.36 |

Abbreviations: F, female; M, Male; BMI, body mass index; SD, standard deviation

**Table 2.**
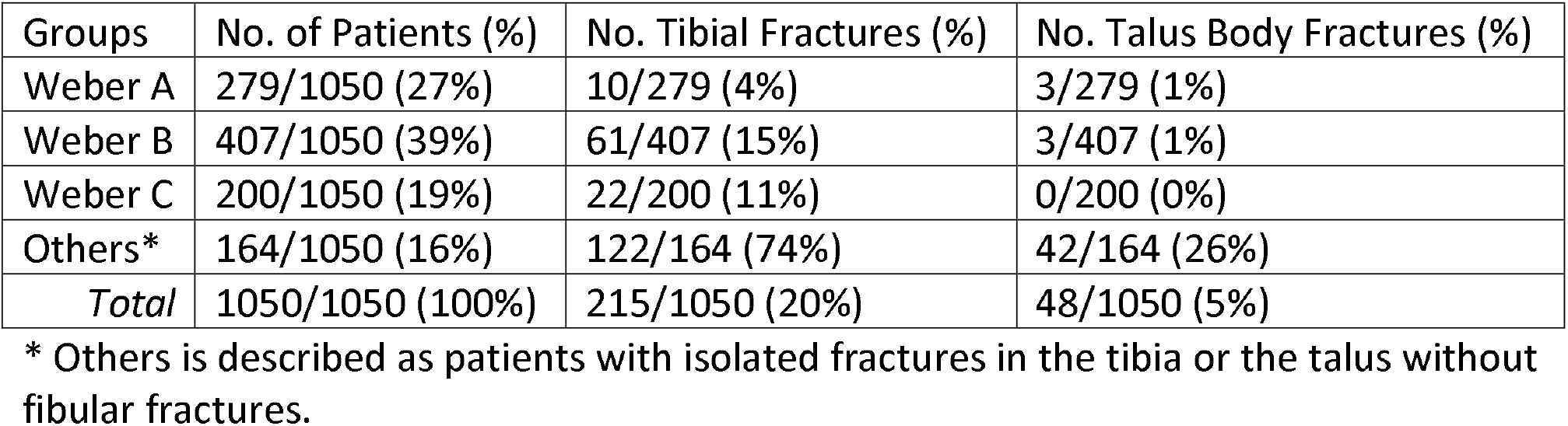
The proportion of ankle fractures based on Danis-Weber classification in a cohort of 1050 patients.

| Groups | No. of Patients (%) | No. Tibial Fractures (%) | No. Talus Body Fractures (%) |
| --- | --- | --- | --- |
| Weber A | 279/1050 (27%) | 10/279 (4%) | 3/279 (1%) |
| Weber B | 407/1050 (39%) | 61/407 (15%) | 3/407 (1%) |
| Weber C | 200/1050 (19%) | 22/200 (11%) | 0/200 (0%) |
| Others* | 164/1050 (16%) | 122/164 (74%) | 42/164 (26%) |
| *Total* | 1050/1050 (100%) | 215/1050 (20%) | 48/1050 (5%) |

\* Others is described as patients with isolated fractures in the tibia or the talus without fibular fractures.

The values evaluating the accuracy of the deep learning algorithms and DCNNs are shown in Table 3. Both Inception V3 model and the Resnet 50 model showed higher accuracy using 3-view radiographic image stacks compared to single-views. Sensitivity, specificity, PPV, NPV, accuracy, F-score, and AUC, were slightly higher in Inception V3 model compared to Resnet 50 (Table 3).

**Table 3.** Performance of two DCNN models in detection of ankle fractures. Single-view (anteroposterior) and three-view radiographs were used to assess the accuracy of each DCNN model.

| Model | No. of Radiographic Views | Sensitivity | Specificity | PPV | NPV | Accuracy | F-Score | AUC |
| --- | --- | --- | --- | --- | --- | --- | --- | --- |
| **Inception V3** | Single view | 90.5% | 93.9% | 93.7% | 91.1% | 92.3% | 92.1% | 95.1% |
|  | Three views | 98.7% | 98.6% | 98.7% | 98.7% | 98.6% | 98.7% | 99.3% |
| **Resnet 50** | Single view | 93.9% | 89.2% | 89.5% | 93.7% | 91.7% | 91.7% | 94.3% |
|  | Three views | 97.5% | 93.9% | 95.2% | 96.8% | 95.9% | 96.3% | 97.8% |

Abbreviations: DCNN, deep convolutional neural networks, PPV, positive predictive value; NPV, negative predictive value; AUC, area under the curve

Evaluating the performance of both models on 72 individuals with occult ankle fractures showed that Inception V3 model could detect 71/72 (98.6%) while Resnet 50 detected 69/72 (95.8%) of the fractures. Using saliency map in our models, the locations of the fractures were highlighted that indicates the accurate decision-making process of our algorithms (Figure 3).

**Figure 3.**
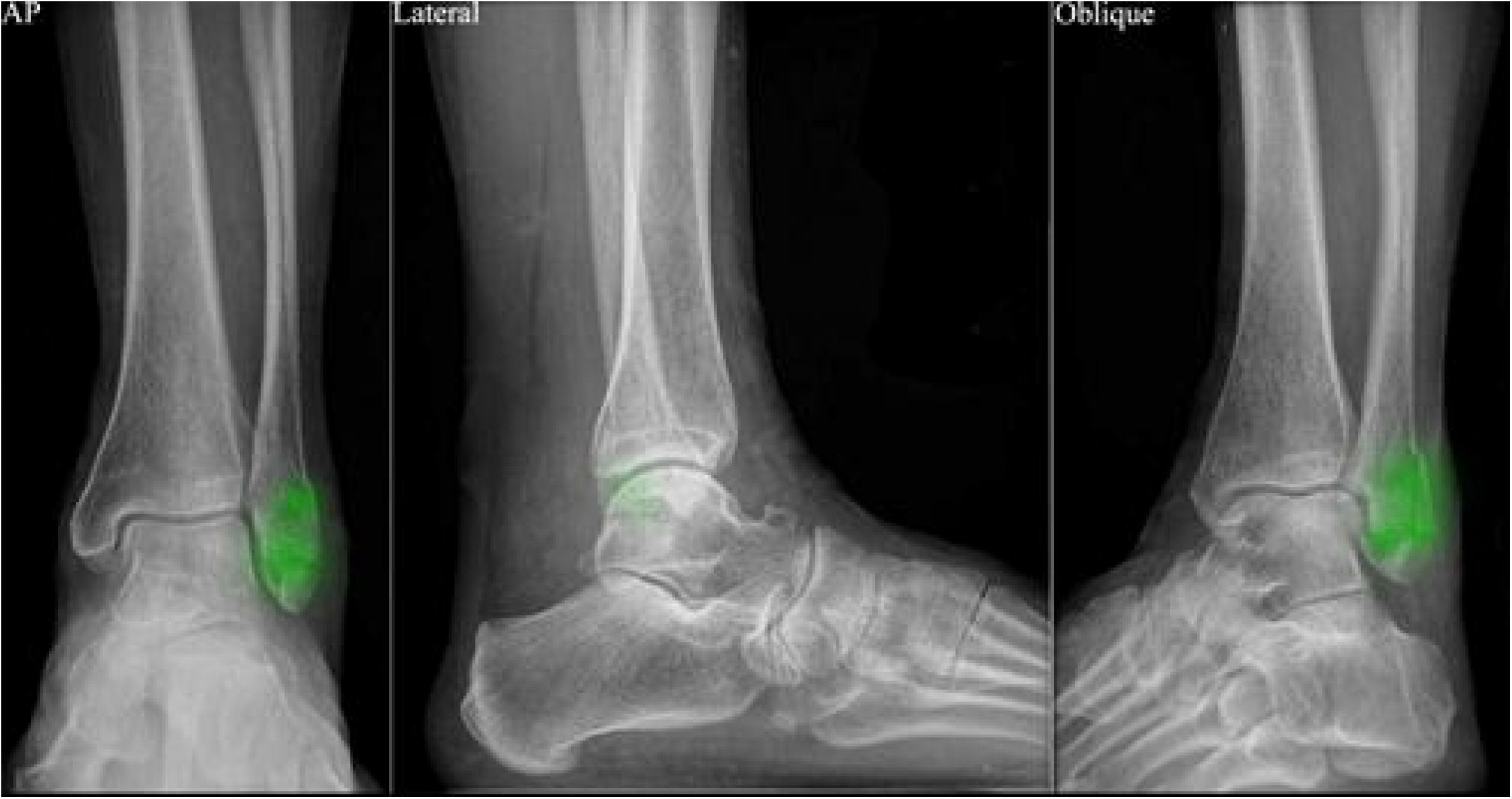
Saliency maps for an ankle radiograph of a patient with lateral malleolus fracture using our deep learning algorithms. The bright green color annotates the location of the fracture. Higher brightness shows the most influential area on the deep convolutional neural network’s performance.

## Discussion

Ankle fractures are one of the most common orthopaedic conditions for which patients are referred to the emergency department^1^. Depending on different factors including knowledge, experience, the burden of patients, time and costs, and quality of the images, the accuracy of assessment varies among healthcare providers ^7, 27^. Hence, while education among care providers remains a high priority, providing computer-assisted image interpretation systems may help their capabilities in recognizing ankle fractures and benefit early decision-making processes for treatment ^8^. The present study shows that using DL and DCNNs for the detection of ankle fractures can lead to diagnostic accuracy of more than 98% using conventional radiographs. Moreover, we found that providing three-view radiographs led to greater performance compared to a single-view radiograph.

In this study, we evaluated the performance of two pretrained DCNNs for the detection of ankle fractures, Inception V3 and Resnet 50. The outcome from Inception V3 neural network outperformed Resnet 50 in overall performance criteria including specificity, sensitivity, PPV, NPV, accuracy, F-score, and AUC, and most importantly in the detection of occult fractures (98.6% vs. 95.8%, respectively). In a similar study on ankle fractures, Kitamura et al. used different pretrained DCNNs including Inception V3, Resnet, and Xception ^28^. They used imaging data from a total of 298 normal and 298 fractured ankles and compared the XX performance of their algorithms using single-view versus three-views. The highest accuracy (83%) belonged to a set of DCNNs being trained using three-view radiographs (vs. 76% by single-view). The authors stated that the accuracy of their method could be improved if they had a larger dataset and if a more appropriate DCNN layer adjustment would be used based on the extracted features from the radiographs and expected outcomes. In this context, a study by Kim et al. using lateral wrist radiographs to re-train Inception V3 DCNN showed an AUC of 0.95, which indicated a satisfactory performance of this network in the detection of fractures of the upper extremity ^29^. In yet another study, Chung et al. found an AUC of 0.94 and an accuracy of 77% for proximal humerus fractures using a pre-trained Resnet DCNN ^30^. Their study comprised 1,891 single-view shoulder radiographs, 1,376 patients, and 515 normal shoulders. One of the reasons for their lower accuracy compared to our study can be the discrepancy between the number of normal and abnormal radiographs. Another reason might be related to the manipulation of the training set, as they cropped the images and resized them to 256 × 256 pixels. This may have reduced the quality of the images and limit the number of extracted features. In our method, we did not use any preprocessing method on our images, which helps to maintain the quality of the image and brings about a more accurate assessment while it would also require higher processing power. Overall, there are no definite criteria to classify the superiority of one algorithm over another be it Inception V3, Resnet, or VGG-16. ^27^. Each algorithm requires to be tested by relatively similar data to the training set and the intermediate layers should be modified based on the desired outcome. For example, if an algorithm is trained by radiographs, it should be tested and validated using radiographs. Moreover, if the algorithm is trained to detect ankle fractures or ankle instability, it should be validated using radiographs obtained from the ankle joint. To modify the DCNN in order to deliver the desired outcome, annotating the radiographs can be helpful. For instance, annotating the fracture site during the training phase, will help the DCNN focus more on this specific type of abnormality in the radiographs. The performance of DCNNs not only depends on the structure of the network, but also relies on the quality and quantity of the training data. If the layers of the DCNN are modified appropriately according to the type of input data, and if the training set is a valid and reliable representative of the actual images of the patients seen in the clinic, the performance of the algorithm can increase significantly.

Occult fractures can be easily missed by the care providers, especially if they have insufficient knowledge or experience recognizing the injury, or due to high workload. Misdiagnosis may lead to degenerative changes of the injury side over time and patient morbidity. Thus, finding a method to help detect ankle fractures faster and more precisely can be valuable. Among the patients included in this study, 72 were suffering from occult fractures that were missed on initial radiographs but were detected in secondary radiographs or CT scans. Our DCNNs showed promising performance in detecting occult fractures with only one missed injury (1/72; 1.4%) using Inception V3 and three missed cases via Resnet 50 (3/72; 4.2%). This shows that a DCNN that is trained using a large population of normal versus abnormal images, is highly unlikely to miss even subtle and occult fractures although these numbers were relatively limited. Urakawa et al. compared the performance of a simple pretrained DCNN, VGG-16 versus orthopaedic surgeons in detecting hip fractures on AP views from 1773 patients ^31^. Their algorithm showed an AUC of 0.98 compared to five surgeons with an AUC of 0.97. One of the strengths of their study was the high number of included patients. A limitation was that in their research data were preprocessed by manipulating the size of the images, contrast, color, orientation, and resolution. This may make the outcome of their algorithm less reliable since the algorithm was not built on original data and testing and validating it on original data obtained from the imaging device may not produce similar results and accuracy. Olczak et al. reported an accuracy of 83% using DCNN compared to 82% obtained from two orthopaedic surgeons in detecting wrist, hand, and ankle fractures using radiographs ^14^. The performance of a human interpreter relies on different factors including knowledge, expertise, time, quality of images, and the viewing software. Despite our hospital being a level 1 trauma center, about 7% of the ankle fractures were missed on primary evaluation. These numbers might be even higher in limited resource settings or in care facilities with relatively inexperienced care-providers. Using DL methods aggregated into X-ray imaging can perform as an assistant, but not as a substitute for the human interpreter and decision-maker aiming to make the hasten the diagnostic process and improving accuracy.

This study has a few limitations that should be taken into consideration. Our data were obtained from a single center. Obtaining data from different centers with different imaging equipment may lead to 1) increased diversity in the images helping our DCNN render more diverse features from images with various qualities, exposure, pixel size, and annotations; 2) increased diversity among included patients, particularly in terms of age and race. This may benefit the accuracy of DCNN when applied to minority subpopulations. Moreover, we trained our model to distinguish images with ankle fractures from normal images. By using a larger dataset, one could develop a DCNN model that can also distinguish various types of ankle fracture.

## Conclusion

The DL algorithms presented in the current study show high accuracy for detecting ankle fractures. Interpretation platforms based on DL methods depicted promising capability to be used assisting care providers in detecting ankle fractures, and potentially other musculoskeletal conditions as well. This may lead to significant improvement in diagnostic accuracy and democratization of knowledge and expertise in imaging interpretation.

## Data Availability

All data referred to in the manuscript are available as per request.

## Acknowledgments

The authors wanted to thank the staff and care providers of the department of orthopaedics and all those who have helped us in performing this study, from designing, to data gathering and preparing the final manuscript.

